# First characterisation of antimicrobial susceptibility and resistance of *Neisseria gonorrhoeae* isolates in Qatar, 2017-2020

**DOI:** 10.1101/2021.10.21.21265124

**Authors:** Muna Al-Maslamani, Emad Bashier Ibrahim Elmagboul, Aslam Puthiyottil, Hiam Chemaitelly, Manoj Kizhakkepeedikayil Varghese, Hamad Eid Al Romaihi, Mohamed H. Al-Thani, Abdullatif Al Khal, Magnus Unemo, Laith J. Abu-Raddad

## Abstract

Limited data are available regarding antimicrobial resistance in *Neisseria gonorrhoeae* strains circulating in WHO Eastern Mediterranean Region (EMR). We investigated the antimicrobial susceptibility/resistance of *N. gonorrhoeae* isolates to five antimicrobials (ceftriaxone, azithromycin, ciprofloxacin, tetracycline, and benzylpenicillin) currently or previously used for gonorrhoea treatment in Qatar, 2017-2020. Minimum inhibitory concentrations (MICs; mg/L) of antimicrobials were determined using Etest on gonococcal isolates collected during January 1, 2017-August 30, 2020 at Hamad Medical Corporation, a national public healthcare provider. During 2017-2020, resistance in isolates from urogenital sites of 433 patients was 64.7% (95% CI: 59.5-69.6%; range: 43.9-78.7%) for ciprofloxacin, 50.7% (95% CI: 45.3-56.1%; range: 41.3-70.4%) for tetracycline, and 30.8% (95% CI: 26.3-35.6%; range: 26.7-35.8%) for benzylpenicillin. Percentage of isolates non-susceptible to azithromycin was 4.1% (95% CI: 2.0-7.4%; range: 2.7-4.8%) and all (100%) isolates were susceptible to ceftriaxone. Two (1.6%) isolates from 2019 and one (2.2%) isolate from 2020 had high-level resistance to azithromycin (MIC≥256 mg/L). Overall, 1.0% (4/418) of isolates had a ceftriaxone MIC of 0.25 mg/L, which is at the ceftriaxone susceptibility breakpoint (MIC≤0.25 mg/L). Treatment with ceftriaxone 250 mg plus azithromycin 1 g can continuously be recommended for gonorrhoea therapy in Qatar. Continued quality-assured gonococcal AMR surveillance is warranted in EMR.

## INTRODUCTION

Gonorrhoea is a common sexually transmitted infection (STI) caused by the bacterium, *Neisseria gonorrhoeae* (gonococcus) [1, 2]. Infection by *N. gonorrhoeae* is associated with cervicitis, pelvic inflammatory disease, ectopic pregnancy, and infertility in women [3-5] and urethritis, epididymitis, and infertility (rare) in men, as well as extragenital infections in both genders [3, 6-8]. In 2016, the World Health Organization (WHO) estimated 87 million new global urogenital gonorrhoea cases each year [9], and WHO has called for achieving a 90% reduction in gonorrhoea incidence by 2030 [10].

Management and control of gonorrhoea have been substantially compromised by widespread antimicrobial resistance (AMR), including international spread of multidrug-resistant and extensively drug-resistant strains with resistance to the last-line, extended-spectrum cephalosporins (ESCs) [2, 11-13]. Resistance had emerged previously to sulphonamides, penicillins, tetracyclines, macrolides, fluoroquinolones, and early-generation cephalosporins [11, 14-16]. In 2012, WHO launched a global action plan to control the spread and impact of AMR in *N. gonorrhoeae* [16], and in 2017 WHO declared gonococcal AMR as a global high priority [17].

One key priority of the WHO global action plan is to enhance gonococcal AMR surveillance globally. Consequently, the WHO’s Global Gonococcal Antimicrobial Surveillance Programme (GASP), initially stabled in 1990[18], was revitalized and strengthened [11, 13]. WHO GASP is identifying emerging AMR, monitoring AMR trends, and informing refinements of global, regional, and national gonorrhoea treatment guidelines, as well as public health strategies and policies developed by WHO and other organisations [11]. WHO recommends that treatment guidelines be refined based on data from recent and quality-assured gonococcal AMR surveillance and that use of an antimicrobial in empiric treatment is be discontinued when the rate of therapeutic failures and/or AMR reach 5% [11, 13, 16, 19]. However, exceedingly limited data regarding gonococcal AMR have been available in several WHO regions, especially in the WHO African and Eastern Mediterranean Regions (‘Arab World’, including the Middle East and North Africa) [11, 13].

Qatar is an Arab country located in the Middle East with a resident population of 2.8 million people [20]. However, 89% of the population are expatriates from over 150 countries [21, 22]. About 60% of the population consists of expatriate craft and manual workers, typically working in mega-development projects [23], who are predominantly young (20-49 years of age), male, and single [24]. The extensive multi-national diversity of this population and its frequent travel between country of origin and Qatar may yield an unusual epidemiological situation for transmission of *N. gonorrhoeae* strains originating from different parts of the world. Thus Qatar provides a unique setting for gonococcal AMR surveillance.

The objectives of this study were to initiate a gonococcal AMR surveillance programme, quality assured in accordance with WHO standards in Qatar, and to investigate the antimicrobial susceptibility of *N. gonorrhoeae* isolates in this country, thereby informing the national STI treatment guidelines in Qatar, as well as contributing to the WHO GASP [11, 13] at a critical time for gonococcal AMR surveillance. With the very limited gonococcal AMR surveillance to date in the WHO Eastern Mediterranean Region [11, 13], this study makes the largest contribution to this surveillance effort from a region that constitutes about 10% of the global population [25].

## MATERIAL AND METHODS

### Study population

All consecutive viable gonococcal isolates cultured from January 1, 2017 to August 30, 2020 at the Microbiology Laboratory at Hamad Medical Corporation (HMC), a national public healthcare provider in Qatar, were included in the study. Gonococcal isolates (n=433) were from 433 patients (or episodes of infection) with urogenital gonorrhoea. Demographic data (nationality, gender, and age) were collected, but no personal identifiers were available in the study. All patients were aimed to be treated in accordance with the national STI Screening and Prevention Guidelines that implement the United States Centers for Disease Control and Prevention (CDC) Sexually Transmitted Diseases Treatment Guidelines [26].

Urethral swab specimens from males and cervical swab specimens from females were collected in Amies transportation medium (International for Medical Equipment and Supplies, Doha, Qatar).

### Culture of *Neisseria gonorrhoeae*

All swabs were transported in ≤12 h to the laboratory and inoculated on selective chocolate culture medium (International for Medical Equipment and Supplies, Doha, Qatar), followed by incubation in 5±1% CO2-enriched humid atmosphere at 36±1°C for 24 h, and if negative, for an additional 24 h. Suspected colonies were verified as *N. gonorrhoeae* using an oxidase test, Bruker Maldi Biotyper (Bruker Daltonik, Bremen, Germany), and Phadebact Monoclonal GC Test (MKL Diagnostic AB, Sollentuna, Sweden). Isolates were stored in a Microbank cryovial system that incorporates 25 treated beads and a proprietary cryopreservative (Pro-Lab Diagnostics, Cheshire, UK) at -80°C.

### Antimicrobial susceptibility testing

Minimum inhibitory concentrations (MICs; mg/L) of five antimicrobials (ceftriaxone, azithromycin, ciprofloxacin, tetracycline, and benzylpenicillin) were determined using Etest (bioMerieux, Marcy-l’Etoile, France), in accordance with the manufacturer’s instructions. Azithromycin was only tested in 2018-2020 and some isolates were not tested against all antimicrobials, i.e. because they were not requested in routine diagnostics, because of a temporary lack of reagents, such as Etest strips, and/or because the isolates were not viable for subsequent antimicrobial susceptibility testing. MIC values were reported and interpreted using whole MIC doubling dilutions, an, where available, clinical breakpoints for susceptibility (S) and resistance (R), according to the United States Clinical Laboratory and Standards Institute (CLSI) [27]. For ceftriaxone and azithromycin, no clinical resistance breakpoints exist and only the recommended susceptibility breakpoints could be used [27]. β-lactamase production was identified using BD BBL Cefinase paper discs (Becton, Dickinson and Company, Franklin Lakes, NJ, USA). The *N. gonorrhoeae* ATCC 49226 international reference strain was used for quality control. The STROBE checklist can be found in Table S1 of Supplementary Information.

## RESULTS

### Patients and characteristics of *Neisseria gonorrhoeae* isolates

*N. gonorrhoeae* isolates (one per patient or gonorrhoea episode) from urogenital sites of 433 patients were examined; 418 (96.5%) were from males and 15 (3.5%) were from females. The patients represented 32 nationalities, but most patients were Qatari (67.2%). All other nationalities each accounted for <10% of the isolates. The median age for the males was 24.0 years (mean: 26.0 years, range: 13-77 years) and for the females it was 32.0 years (mean: 36.8 years, range: 24-60 years).

### Antimicrobial susceptibility of *Neisseria gonorrhoeae* isolates

Results of antimicrobial susceptibility testing of all *N. gonorrhoeae* isolates (n=433) are listed in Table S2 of Supplementary Information and are summarised in Table 1.

**Table 1.** Antimicrobial susceptibility in *Neisseria gonorrhoeae* isolates (n=433) from Qatar, 2017-2020

| Antimicrobials<br>(No. tested) | Susceptible/Intermediate/Resistant (/Non-susceptible), %<sup>a</sup> |  |  |  |  |
| --- | --- | --- | --- | --- | --- |
|  | 2017<br>(n=109) | 2018<br>(n=152) | 2019<br>(n=126) | 2020<br>(n=46) | Total<br>(n=433) |
| CRO (419) | 100/NA/0 | 100/NA/0 | 100/NA/0 | 100/NA/0 | 100/NA/0 |
| AZM (234) | - | 97.3/NA/2.7 | 95.2/NA/4.8 | 95.7/NA/4.3 | 95.9/NA/4.1 |
| CIP (365) | 7.5/13.8/78.7 | 11.6/25.9/62.6 | 13.3/24.1/62.7 | 19.5/36.6/43.9 | 11.8/23.6/64.7 |
| PEN (399) | 12.8/51.4/35.8 | 14.8/58.5/26.7 | 14.7/53.2/32.1 | 19.6/52.2/28.3 | 14.8/54.4/30.8 |
| TET (345) | 8.6/21.0/70.4 | 18.8/36.1/45.1 | 17.6/36.5/46.0 | 23.9/34.8/41.3 | 16.8/32.5/50.7 |

No., number; CRO, ceftriaxone; AZM, azithromycin; CIP, ciprofloxacin; PEN, benzylpenicillin; TET, tetracycline; NA, not applicable; –, not tested.
<sup>a</sup>The breakpoints (susceptible, resistant) were as follows: ceftriaxone (MIC≤0.25 mg/L, not available), azithromycin (MIC≤1 mg/L, not available), ciprofloxacin (MIC≤0.06 mg/L, MIC≥1 mg/L), benzylpenicillin (MIC≤0.06 mg/L, MIC≥2.0 mg/L) and tetracycline (MIC≤0.25 mg/L, MIC≥2 mg/L).

Overall, in 2017-2020, resistance to ciprofloxacin, tetracycline, and benzylpenicillin was 64.7% (95% confidence interval (CI): 59.5-69.6%; range during 2017-2020: 43.9-78.7%), 50.7% (95%CI: 45.3-56.1%; range: 41.3-70.4%), and 30.8% (95% CI: 26.3-35.6%; range: 26.7-35.8%), respectively. The percentage of isolates non-susceptible to azithromycin was 4.1% (95% CI: 2.0-7.4%; range: 2.7-4.8%) and all (100%) isolates were susceptible to ceftriaxone (Table 1).

MIC distributions for ceftriaxone and azithromycin, included in the internationally recommended first-line, dual antimicrobial therapies [2, 14], are presented in Fig. 1. In total, 1.0% (4/418) of isolates had a ceftriaxone MIC of 0.25 mg/L, which is at the ceftriaxone susceptibility breakpoint [MIC≤0.25 mg/L [27]]: 1.9% (2/108) in 2017, 0.9% (1/113) in 2019, and 2.2% (1/46) in 2020. The percentage of isolates with ceftriaxone MIC≤0.016 mg/L was 86.6%. Most isolates (>95% having a MIC of <2 mg/L) belonged to the azithromycin MIC wild-type distribution (Fig. 1). Notably, two (1.6%) isolates from 2019 and one (2.2%) isolate from 2020 had a high-level resistance to azithromycin (MIC≥256 mg/L).

## DISCUSSION

AMR in *N. gonorrhoeae* remains a global public health concern, compromising management and control of gonorrhoea, and enhanced, quality-assured gonococcal AMR surveillance internationally is imperative. However, in the most recently published WHO GASP data (from 2015-2016) only one (4.8%) member state (Pakistan) in the WHO Eastern Mediterranean Region provided AMR data, and only 64 gonococcal isolates were examined [13]. In the present study, we substantially increased the available gonococcal AMR data from this WHO region, and provided AMR data for 433 isolates (five antimicrobials) cultured in Qatar in 2017-2020, which will be available for the WHO GASP. All antimicrobial susceptibility testing was also based on MIC determination, that is, Etest was used, which is recommended in the WHO GASP [11, 13].

High levels of resistance to ciprofloxacin, tetracycline, and benzylpenicillin were found. It is notable that resistance to ciprofloxacin was very high at 64.7%, which is similar to the high levels reported in several other WHO regions [11, 13]. Despite high levels of ciprofloxacin resistance, this antimicrobial or other fluoroquinolones remain in use for empirical treatment of gonorrhoea in many countries. This use needs to be abandoned and ciprofloxacin should only be used for treatment of gonorrhoea when the infection has been proven ciprofloxacin-susceptible as a result of laboratory testing [28].

No resistance to ceftriaxone was found among isolates in Qatar in 2017-2020 and only a small proportion (4.1%) of isolates demonstrated resistance to azithromycin. Accordingly, current first-line empiric treatment in Qatar, using ceftriaxone and azithromycin based on the United States CDC Sexually Transmitted Diseases Treatment Guidelines [26], remains an appropriate treatment, which additionally eradicates concomitant *Chlamydia trachomatis* infection and many *Mycoplasma genitalium* infections. Nevertheless, it is concerning that two isolates from 2019 and one isolate from 2020 in Qatar had high-level resistance to azithromycin (MIC≥256 mg/L). Furthermore, recently published new recommendations for gonorrhoea treatment (ceftriaxone 500 mg monotherapy) in the United States CDC Sexually Transmitted Diseases Treatment Guidelines [29] will be considered.

The limitations of the present study included the lack of detailed epidemiological information of all gonorrhoea cases and uncertainties regarding the representativeness of the examined gonorrhoea cases. Individuals diagnosed with gonorrhoea at HMC, a national public healthcare provider, were examined, but we did not access any cases being managed in the private sector or at other public healthcare facilities, such as the Qatar Red Crescent Society, the main healthcare provider for craft and manual workers. Nevertheless, 32 nationalities were represented, which suggests that some expatriate craft and manual workers were represented. Only urogenital samples were collected. Azithromycin susceptibility testing was not conducted prior to 2018 and some of the isolates were not examined for susceptibility to all antimicrobials, because they were not requested in routine diagnostics, because of temporary lack of reagents, such as Etest strips, and/or because the isolates were not viable for subsequent antimicrobial susceptibility testing. Finally, the 2016 WHO *N. gonorrhoeae* reference strains [30] were not used for quality assurance and control; however, these strains are now available for use in AMR surveillance.

In conclusion, high levels of resistance to ciprofloxacin, tetracycline, and benzylpenicillin were found in Qatar, but less resistance to azithromycin and none to ceftriaxone. Treatment with ceftriaxone 250 mg plus azithromycin 1 g can be recommended for gonorrhoea therapy in Qatar, but the recently recommended ceftriaxone 500 mg monotherapy [29] will also be considered. Continued and enhanced quality-assured gonococcal AMR surveillance is essential in Qatar and other countries in the WHO Eastern Mediterranean Region. Whole-genome sequencing of *N. gonorrhoeae* isolates is also in progress to enhance understanding of *N. gonorrhoeae* AMR determinants and transmission of *N. gonorrhoeae*, as well as its resistance in different subpopulations in Qatar, compared with the international population.

## Data Availability

All relevant data are available within the manuscript and the supplementary material.

## Acknowledgements

The authors are grateful for the administrative support of Ms. Adona Canlas.

## Contributors

LJA, MU, MA-M, and EBE conceived and designed the study. EBE and AP conducted the laboratory work and antimicrobial susceptibility testing. MU and HC conducted the statistical analyses. MU and LJA with MA-M, EBE, and HC wrote the first draft of the article. All authors contributed to data collection and acquisition, database development, discussion and interpretation of the results, and to the writing of the manuscript. All authors have read and approved the final manuscript.

## Patient consent for publication

Not required.

## Ethical approval

The study was approved by the Weill Cornell Medicine - Qatar and Hamad Medical Corporation Institutional Review Boards. The study was conducted following the ethics review boards guidelines and regulations. Waiver of consenting was granted as per the study design, retrospective collection of deidentified gonococcal isolates.

## Data availability statement

All relevant data are available within the manuscript and the supplementary material.

## Supplementary information

**Table S1.** STROBE checklist of items that should be included in reports of cross-sectional studies

|  | Item No | Recommendation | Page No |
| --- | --- | --- | --- |
| Title and abstract | 1 | (a) Indicate the study’s design with a commonly used term in the title or the abstract | 3 |
|  |  | (b) Provide in the abstract an informative and balanced summary of what was done and what was found | 3 |
| Introduction |  |  |  |
| Background/rationale | 2 | Explain the scientific background and rationale for the investigation being reported | 5-6 |
| Objectives | 3 | State specific objectives, including any prespecified hypotheses | 6 |
| Methods |  |  |  |
| Study design | 4 | Present key elements of study design early in the paper | 7 |
| Setting | 5 | Describe the setting, locations, and relevant dates, including periods of recruitment, exposure, follow-up, and data collection | 7 |
| Participants | 6 | (a) Give the eligibility criteria, and the sources and methods of selection of participants | 7 |
| Variables | 7 | Clearly define all outcomes, exposures, predictors, potential confounders, and effect modifiers. Give diagnostic criteria, if applicable | 7-8 |
| Data sources/ measurement | 8* | For each variable of interest, give sources of data and details of methods of assessment (measurement). Describe comparability of assessment methods if there is more than one group | 7-8 |
| Bias | 9 | Describe any efforts to address potential sources of bias | 8 |
| Study size | 10 | Explain how the study size was arrived at | 7 |
| Quantitative variables | 11 | Explain how quantitative variables were handled in the analyses. If applicable, describe which groupings were chosen and why | 8 |
| Statistical methods | 12 | (a) Describe all statistical methods, including those used to control for confounding | 8 |
|  |  | (b) Describe any methods used to examine subgroups and interactions | 8 |
|  |  | (c) Explain how missing data were addressed | 8 |
|  |  | (d) If applicable, describe analytical methods taking account of sampling strategy | 8 |
|  |  | (e) Describe any sensitivity analyses | NA |
| Results |  |  |  |
| Participants | 13* | (a) Report numbers of individuals at each stage of study—eg numbers potentially eligible, examined for eligibility, confirmed eligible, included in the study, completing follow-up, and analysed | 8 |
|  |  | (b) Give reasons for non-participation at each stage | NA |
|  |  | (c) Consider use of a flow diagram | NA |
| Descriptive data | 14* | (a) Give characteristics of study participants (eg demographic, clinical, social) and information on exposures and potential confounders | 8-9 |
|  |  | (b) Indicate number of participants with missing data for each variable of interest | Table S1 in Supplementary Material |
| Outcome data | 15* | Report numbers of outcome events or summary measures | 9, Table 1 and Figure 1 |
| Main results | 16 | (a) Give unadjusted estimates and, if applicable, confounder-adjusted estimates and their precision (eg, 95% confidence interval). Make clear which confounders were adjusted for and why they were included | 9 |
|  |  | (b) Report category boundaries when continuous variables were categorized | 9, Table 1 and Figure 1 |
|  |  | (c) If relevant, consider translating estimates of relative risk into absolute risk for a meaningful time period | NA |
| Other analyses | 17 | Report other analyses done—eg analyses of subgroups and interactions, and sensitivity analyses | 9, Table 1 and Figure 1 |
| Discussion |  |  |  |
| Key results | 18 | Summarise key results with reference to study objectives | 9-10 |
| Limitations | 19 | Discuss limitations of the study, taking into account sources of potential bias or imprecision. Discuss both direction and magnitude of any potential bias | 11 |
| Interpretation | 20 | Give a cautious overall interpretation of results considering objectives, limitations, multiplicity of analyses, results from similar studies, and other relevant evidence | 9-11 |
| Generalisability | 21 | Discuss the generalisability (external validity) of the study results | 11 |
| **Other information** |  |  |  |
| Funding | 22 | Give the source of funding and the role of the funders for the present study and, if applicable, for the original study on which the present article is based | 15 |

NA, not applicable.

**Table S2.** Patient characteristics and antimicrobial susceptibility in *Neisseria gonorrhoeae* isolates (n=433) from Qatar, 2017-2020

| Isolate | Year | Sex | Age (years) | Benzylpenicillin |  | Ceftriaxone |  | Ciprofloxacin |  | Tetracycline |  | Azithromycin |  |
| --- | --- | --- | --- | --- | --- | --- | --- | --- | --- | --- | --- | --- | --- |
|  |  |  |  | MIC | Result | MIC | Result | MIC | Result | MIC | Result | MIC | Result |
| 1 | 2017 | M | 20-24 | 1 | I | 0.032 | S | 8 | R | - | - | - | - |
| 2 | 2017 | M | 45-49 | 0.38 | I | 0.012 | S | 0.38 | I | - | - | - | - |
| 3 | 2017 | M | 40-44 | 0.032 | S | 0.002 | S | 16 | R | 0.19 | S | - | - |
| 4 | 2017 | M | 20-24 | 1.5 | R | 0.032 | S | 4 | R | 4 | R | - | - |
| 5 | 2017 | M | 20-24 | 0.094 | I | 0.003 | S | 4 | R | - | - | - | - |
| 6 | 2017 | M | 20-24 | 256 | R | 0.006 | S | 2 | R | 3 | R | - | - |
| 7 | 2017 | M | 35-39 | 8 | R | 0.008 | S | 32 | R | - | - | - | - |
| 8 | 2017 | M | 25-29 | 0.064 | S | 0.004 | S | 4 | R | 1.5 | R | - | - |
| 9 | 2017 | M | 25-29 | 0.25 | I | 0.004 | S | 0.75 | R | 24 | R | - | - |
| 10 | 2017 | M | 30-34 | 0.25 | I | 0.003 | S | 8 | R | 24 | R | - | - |
| 11 | 2017 | F | 30-34 | 1 | I | 0.003 | S | 0.5 | I | 16 | R | - | - |
| 12 | 2017 | M | 20-24 | 0.75 | I | 0.023 | S | 8 | R | - | - | - | - |
| 13 | 2017 | M | 30-34 | 1.5 | R | 0.012 | S | 32 | R | 3 | R | - | - |
| 14 | 2017 | M | 30-34 | 0.38 | I | 0.023 | S | 1.5 | R | 1.5 | R | - | - |
| 15 | 2017 | M | 30-34 | 12 | R | 0.064 | S | 1 | R | - | - | - | - |
| 16 | 2017 | M | 20-24 | 0.75 | I | 0.023 | S | 8 | R | - | - | - | - |
| 17 | 2017 | M | 15-19 | 16 | R | 0.004 | S | 1 | R | - | - | - | - |
| 18 | 2017 | M | 25-29 | 12 | R | 0.003 | S | 1.5 | R | - | - | - | - |
| 19 | 2017 | M | 20-24 | 0.5 | I | 0.012 | S | 6 | R | - | - | - | - |
| 20 | 2017 | M | 15-19 | 8 | R | 0.002 | S | 2 | R | - | - | - | - |
| 21 | 2017 | M | 25-29 | 0.25 | I | 0.006 | S | 2 | R | - | - | - | - |
| 22 | 2017 | M | 20-24 | 0.125 | I | 0.012 | S | 3 | R | - | - | - | - |
| 23 | 2017 | M | 25-29 | 0.19 | I | 0.032 | S | 8 | R | - | - | - | - |
| 24 | 2017 | M | 25-29 | 0.75 | I | 0.002 | S | 8 | R | - | - | - | - |
| 25 | 2017 | M | 30-34 | 0.016 | S | 0.002 | S | 2 | R | - | - | - | - |
| 26 | 2017 | M | 25-29 | 0.125 | I | 0.004 | S | 0.006 | S | - | - | - | - |
| 27 | 2017 | M | 20-24 | 0.19 | I | 0.012 | S | 4 | R | - | - | - | - |
| 28 | 2017 | M | 25-29 | 0.38 | I | 0.008 | S | 2 | R | - | - | - | - |
| 29 | 2017 | M | 35-39 | 32 | R | 0.004 | S | 1 | R | - | - | - | - |
| 30 | 2017 | M | 20-24 | 8 | R | 0.006 | S | 1.5 | R | - | - | - | - |
| 31 | 2017 | M | 20-24 | 0.064 | S | 0.006 | S | 32 | R | - | - | - | - |
| 32 | 2017 | M | 20-24 | 0.75 | I | 0.012 | S | 6 | R | 1.5 | R | - | - |
| 33 | 2017 | M | 25-29 | 6 | R | 0.004 | S | 1.5 | R | 16 | R | - | - |
| 34 | 2017 | F | 25-29 | 4 | R | 0.003 | S | 1 | R | 16 | R | - | - |
| 35 | 2017 | F | 50-54 | 0.19 | I | 0.023 | S | 3 | R | 32 | R | - | - |
| 36 | 2017 | M | 25-29 | 0.38 | I | 0.016 | S | 32 | R | - | - | - | - |
| 37 | 2017 | M | 15-19 | 12 | R | 0.003 | S | 1 | R | 1 | I | - | - |
| 38 | 2017 | M | 25-29 | 0.5 | I | 0.012 | S | 32 | R | 2 | R | - | - |
| 39 | 2017 | M | 25-29 | 0.125 | I | 0.002 | S | 4 | R | 1.5 | R | - | - |
| 40 | 2017 | M | 15-19 | 3 | R | 0.002 | S | 1.5 | R | 8 | R | - | - |
| 41 | 2017 | M | 25-29 | 32 | R | -* | S* | 4 | R | 0.5 | I | - | - |
| 42 | 2017 | M | 30-34 | 0.016 | S | 0.002 | S | - | - | 16 | R | - | - |
| 43 | 2017 | M | 20-24 | 0.064 | S | 0.003 | S | 0.002 | S | 0.25 | S | - | - |
| 44 | 2017 | M | 35-39 | 192 | R | 0.003 | S | 2 | R | 24 | R | - | - |
| 45 | 2017 | M | 20-24 | 3 | R | 0.003 | S | 1.5 | R | 12 | R | - | - |
| 46 | 2017 | M | 25-29 | 0.016 | S | 0.002 | S | 0.016 | S | 1 | I | - | - |
| 47 | 2017 | M | 20-24 | 16 | R | 0.003 | S | 3 | R | 6 | R | - | - |
| 48 | 2017 | M | 20-24 | 0.016 | S | 0.003 | S | 32 | R | 0.25 | S | - | - |
| 49 | 2017 | M | 35-39 | 0.25 | I | 0.003 | S | 0.75 | R | 32 | R | - | - |
| 50 | 2017 | M | 15-19 | 0.38 | I | 0.003 | S | 0.004 | S | 0.5 | I | - | - |
| 51 | 2017 | M | 35-39 | 0.125 | I | 0.012 | S | 8 | R | 1 | I | - | - |
| 52 | 2017 | M | 40-44 | 0.5 | I | 0.023 | S | - | - | 32 | R | - | - |
| 53 | 2017 | M | 30-34 | 0.25 | I | 0.002 | S | - | - | 4 | R | - | - |
| 54 | 2017 | M | 25-29 | 0.38 | I | 0.023 | S | - | - | 1.5 | R | - | - |
| 55 | 2017 | M | 30-34 | 4 | R | 0.004 | S | - | - | 16 | R | - | - |
| 56 | 2017 | M | 15-19 | 0.25 | I | 0.012 | S | - | - | 1.5 | R | - | - |
| 57 | 2017 | M | 30-34 | 0.125 | I | 0.004 | S | - | - | 0.38 | I | - | - |
| 58 | 2017 | M | 15-19 | 0.25 | I | 0.023 | S | - | - | 1 | I | - | - |
| 59 | 2017 | M | 20-24 | 0.016 | S | 0.002 | S | - | - | 0.25 | S | - | - |
|  |  |  |  | MIC | Result | MIC | Result | MIC | Result | MIC | Result | MIC | Result |
| 60 | 2017 | M | 20-24 | 96 | R | 0.003 | S | - | - | 16 | R | - | - |
| 61 | 2017 | M | 20-24 | 0.38 | I | 0.012 | S | - | - | 0.5 | I | - | - |
| 62 | 2017 | M | 20-24 | 1 | I | 0.19 | S | - | - | 3 | R | - | - |
| 63 | 2017 | M | 20-24 | 1 | I | 0.006 | S | 1 | R | 8 | R | - | - |
| 64 | 2017 | F | 25-29 | 0.25 | I | 0.006 | S | 2 | R | 0.25 | S | - | - |
| 65 | 2017 | M | 20-24 | 0.125 | I | 0.016 | S | 1 | R | 0.5 | I | - | - |
| 66 | 2017 | M | 20-24 | 0.047 | S | 0.002 | S | 0.125 | I | 2 | R | - | - |
| 67 | 2017 | M | 20-24 | 0.75 | I | 0.25 | S | 0.047 | S | 0.064 | S | - | - |
| 68 | 2017 | M | 20-24 | 0.5 | I | 0.032 | S | 6 | R | 0.75 | I | - | - |
| 69 | 2017 | M | 15-19 | 16 | R | 0.032 | S | 8 | R | 12 | R | - | - |
| 70 | 2017 | M | 30-34 | 1 | I | 0.032 | S | 6 | R | 2 | R | - | - |
| 71 | 2017 | M | 25-29 | 0.016 | S | 0.002 | S | 4 | R | 0.38 | I | - | - |
| 72 | 2017 | M | 25-29 | 2 | R | 0.003 | S | 8 | R | - | - | - | - |
| 73 | 2017 | M | 15-19 | 0.125 | I | 0.002 | S | 1 | R | 12 | R | - | - |
| 74 | 2017 | M | 30-34 | 0.125 | I | 0.003 | S | 0.5 | I | - | - | - | - |
| 75 | 2017 | M | 15-19 | 8 | R | 0.004 | S | 0.75 | R | 16 | R | - | - |
| 76 | 2017 | M | 15-19 | 2 | R | 0.003 | S | 3 | R | - | - | - | - |
| 77 | 2017 | M | 25-29 | 0.25 | I | 0.016 | S | 2 | R | - | - | - | - |
| 78 | 2017 | M | 15-19 | 0.38 | I | 0.016 | S | 2 | R | 1 | I | - | - |
| 79 | 2017 | M | 20-24 | 4 | R | 0.004 | S | - | - | 32 | R | - | - |
| 80 | 2017 | M | 25-29 | 0.064 | S | 0.003 | S | 0.75 | R | - | - | - | - |
| 81 | 2017 | M | 15-19 | 0.125 | I | 0.004 | S | 0.75 | R | 16 | R | - | - |
| 82 | 2017 | M | 20-24 | 4 | R | 0.004 | S | - | - | 24 | R | - | - |
| 83 | 2017 | M | 25-29 | 8 | R | 0.003 | S | - | - | 12 | R | - | - |
| 84 | 2017 | F | 60-64 | 0.25 | I | 0.003 | S | 0.002 | S | 16 | R | - | - |
| 85 | 2017 | M | 35-39 | 0.064 | S | 0.003 | S | 0.003 | S | 0.5 | I | - | - |
| 86 | 2017 | M | 20-24 | 3 | R | 0.003 | S | 0.5 | I | 12 | R | - | - |
| 87 | 2017 | M | 20-24 | 2 | R | 0.008 | S | 1 | R | 16 | R | - | - |
| 88 | 2017 | M | 15-19 | 0.5 | I | 0.047 | S | 2 | R | 1 | I | - | - |
| 89 | 2017 | M | 20-24 | 0.5 | I | 0.047 | S | 4 | R | 1.5 | R | - | - |
| 90 | 2017 | M | 25-29 | 0.125 | I | 0.003 | S | 0.75 | R | 16 | R | - | - |
| 91 | 2017 | M | 25-29 | 0.094 | I | 0.002 | S | 0.75 | R | 16 | R | - | - |
| 92 | 2017 | M | 15-19 | 6 | R | 0.004 | S | 0.75 | R | 2 | R | - | - |
| 93 | 2017 | M | 25-29 | 0.125 | I | 0.002 | S | 0.75 | R | 0.5 | I | - | - |
| 94 | 2017 | M | 15-19 | 256 | R | 0.004 | S | 0.75 | R | 16 | R | - | - |
| 95 | 2017 | M | 15-19 | 0.38 | I | 0.032 | S | 8 | R | 1 | I | - | - |
| 96 | 2017 | M | 30-34 | 4 | R | 0.002 | S | 0.75 | R | 12 | R | - | - |
| 97 | 2017 | M | 15-19 | 0.064 | S | 0.008 | S | 0.38 | I | 16 | R | - | - |
| 98 | 2017 | M | 15-19 | 2 | R | 0.002 | S | 0.38 | I | 1.5 | R | - | - |
| 99 | 2017 | M | 20-24 | 4 | R | 0.003 | S | 0.38 | I | 6 | R | - | - |
| 100 | 2017 | M | 25-29 | 0.125 | I | 0.004 | S | 0.38 | I | 16 | R | - | - |
| 101 | 2017 | M | 15-19 | 0.75 | I | 0.047 | S | 4 | R | 1.5 | R | - | - |
| 102 | 2017 | M | 20-24 | 8 | R | 0.004 | S | 32 | R | 16 | R | - | - |
| 103 | 2017 | M | 15-19 | 0.125 | I | 0.008 | S | 0.38 | I | 0.25 | S | - | - |
| 104 | 2017 | M | 15-19 | 12 | R | 0.002 | S | 0.25 | I | 12 | R | - | - |
| 105 | 2017 | M | 20-24 | 0.38 | I | 0.008 | S | 1.5 | R | 0.75 | I | - | - |
| 106 | 2017 | M | 20-24 | 8 | R | 0.125 | S | 0.38 | I | 12 | R | - | - |
| 107 | 2017 | M | 15-19 | 8 | R | 0.003 | S | 0.38 | I | 24 | R | - | - |
| 108 | 2017 | M | 15-19 | 0.5 | I | 0.023 | S | 3 | R | 1.5 | R | - | - |
| 109 | 2017 | M | 15-19 | 2 | R | 0.023 | S | 2 | R | 2 | R | - | - |
| 110 | 2018 | M | 30-34 | 4 | R | 0.002 | S | 0.012 | S | 8 | R | - | - |
| 111 | 2018 | M | 25-29 | - | - | 0.003 | S | 0.38 | I | 3 | R | - | - |
| 112 | 2018 | M | 20-24 | 6 | R | 0.003 | S | 0.75 | R | 8 | R | - | - |
| 113 | 2018 | M | 30-34 | 4 | R | 0.002 | S | 0.38 | I | 0.75 | I | - | - |
| 114 | 2018 | M | 15-19 | 0.38 | I | 0.012 | S | 1 | R | 0.5 | I | - | - |
| 115 | 2018 | M | 15-19 | 0.19 | I | 0.002 | S | 0.002 | S | 0.5 | I | - | - |
| 116 | 2018 | M | 25-29 | 0.38 | I | 0.023 | S | 1.5 | R | 0.75 | I | - | - |
| 117 | 2018 | M | 20-24 | 0.38 | I | 0.032 | S | 32 | R | 1 | I | - | - |
| 118 | 2018 | M | 20-24 | 0.19 | I | 0.016 | S | 2 | R | 0.75 | I | - | - |
| 119 | 2018 | M | 15-19 | 0.094 | I | 0.002 | S | 1 | R | 12 | R | - | - |
| 120 | 2018 | M | 30-34 | 0.38 | I | 0.016 | S | 1 | R | 0.75 | I | - | - |
| 121 | 2018 | M | 20-24 | 0.38 | I | 0.006 | S | 0.25 | I | 6 | R | - | - |
| 122 | 2018 | M | 15-19 | 24 | R | 0.004 | S | 0.5 | I | 12 | R | - | - |
|  |  |  |  | MIC | Result | MIC | Result | MIC | Result | MIC | Result | MIC | Result |
| 123 | 2018 | M | 15-19 | 8 | R | 0.004 | S | 0.5 | I | 16 | R | - | - |
| 124 | 2018 | M | 15-19 | - | - | 0.008 | S | 1.5 | R | 0.25 | S | - | - |
| 125 | 2018 | M | 30-34 | - | - | 0.002 | S | 0.002 | S | 6 | R | - | - |
| 126 | 2018 | M | 30-34 | 2 | R | 0.002 | S | 0.19 | I | 0.125 | S | - | - |
| 127 | 2018 | M | 20-24 | 0.38 | I | 0.008 | S | 0.75 | R | 0.25 | S | - | - |
| 128 | 2018 | M | 30-34 | 4 | R | 0.002 | S | 0.75 | R | 6 | R | - | - |
| 129 | 2018 | M | 20-24 | 0.75 | I | 0.032 | S | 4 | R | 1 | I | - | - |
| 130 | 2018 | M | 45-49 | - | - | 0.002 | S | 1.5 | R | 0.38 | I | - | - |
| 131 | 2018 | M | 30-34 | 0.38 | I | 0.016 | S | 2 | R | 1.5 | R | - | - |
| 132 | 2018 | M | 15-19 | 0.38 | I | 0.003 | S | 12 | R | 0.5 | I | - | - |
| 133 | 2018 | M | 25-29 | 0.5 | I | 0.016 | S | 4 | R | 1 | I | - | - |
| 134 | 2018 | M | 15-19 | - | - | 0.003 | S | 0.002 | S | 16 | R | - | - |
| 135 | 2018 | M | 20-24 | 0.75 | I | 0.023 | S | 2 | R | 1 | I | - | - |
| 136 | 2018 | M | 25-29 | 0.064 | S | 0.003 | S | 0.5 | I | 8 | R | - | - |
| 137 | 2018 | M | 15-19 | 16 | R | 0.002 | S | 1 | R | 12 | R | - | - |
| 138 | 2018 | M | 20-24 | - | - | 0.002 | S | 0.38 | I | 16 | R | - | - |
| 139 | 2018 | M | 30-34 | 256 | R | 0.003 | S | 0.19 | I | 8 | R | - | - |
| 140 | 2018 | M | 20-24 | 192 | R | 0.002 | S | 1.5 | R | 16 | R | - | - |
| 141 | 2018 | M | 30-34 | 0.5 | I | 0.023 | S | 2 | R | 0.75 | I | - | - |
| 142 | 2018 | M | 25-29 | 0.25 | I | 0.006 | S | 2 | R | 0.75 | I | - | - |
| 143 | 2018 | M | 25-29 | 0.094 | I | 0.002 | S | 0.002 | S | 24 | R | - | - |
| 144 | 2018 | M | 15-19 | 0.38 | I | 0.008 | S | 0.125 | I | 16 | R | - | - |
| 145 | 2018 | M | 15-19 | 0.38 | I | 0.003 | S | 8 | R | 0.75 | I | - | - |
| 146 | 2018 | M | 25-29 | 0.19 | I | 0.012 | S | 4 | R | 0.75 | I | - | - |
| 147 | 2018 | M | 15-19 | 16 | R | 0.002 | S | 0.38 | I | 12 | R | - | - |
| 148 | 2018 | M | 15-19 | 32 | R | 0.003 | S | 2 | R | 16 | R | - | - |
| 149 | 2018 | M | 45-49 | 3 | R | 0.006 | S | 2 | R | 24 | R | - | - |
| 150 | 2018 | M | 40-44 | 0.016 | S | 0.002 | S | 3 | R | 0.19 | S | - | - |
| 151 | 2018 | M | 30-34 | 0.016 | S | 0.002 | S | 4 | R | 0.25 | S | - | - |
| 152 | 2018 | M | 15-19 | 0.25 | I | 0.008 | S | 2 | R | 0.5 | I | - | - |
| 153 | 2018 | M | 20-24 | 0.016 | S | 0.002 | S | 0.002 | S | 0.19 | S | - | - |
| 154 | 2018 | M | 25-29 | 0.047 | S | 0.002 | S | 0.5 | I | 3 | R | - | - |
| 155 | 2018 | M | 15-19 | 0.5 | I | 0.012 | S | 3 | R | 0.5 | I | - | - |
| 156 | 2018 | M | 25-29 | 16 | R | 0.003 | S | 1 | R | 0.75 | I | - | - |
| 157 | 2018 | M | 20-24 | 0.094 | I | 0.003 | S | 0.5 | I | 8 | R | - | - |
| 158 | 2018 | M | 30-34 | 0.19 | I | 0.012 | S | 4 | R | 1 | I | - | - |
| 159 | 2018 | M | 30-34 | 1 | I | 0.032 | S | 4 | R | 1.5 | R | - | - |
| 160 | 2018 | M | 25-29 | 0.5 | I | 0.023 | S | 6 | R | 1 | I | - | - |
| 161 | 2018 | M | 25-29 | 0.19 | I | 0.012 | S | 1 | R | 0.75 | I | - | - |
| 162 | 2018 | M | 40-44 | 0.19 | I | 0.003 | S | 0.75 | R | 32 | R | - | - |
| 163 | 2018 | M | 30-34 | 16 | R | 0.002 | S | 0.5 | I | - | - | - | - |
| 164 | 2018 | M | 25-29 | 0.75 | I | 0.012 | S | 2 | R | - | - | - | - |
| 165 | 2018 | M | 25-29 | 1 | I | 0.064 | S | 4 | R | - | - | - | - |
| 166 | 2018 | M | 20-24 | 0.19 | I | 0.003 | S | - | - | 0.38 | I | - | - |
| 167 | 2018 | M | 30-34 | 1.5 | R | 0.008 | S | - | - | 1.5 | R | - | - |
| 168 | 2018 | M | 20-24 | 256 | R | 0.004 | S | 2 | R | 2 | R | - | - |
| 169 | 2018 | M | 30-34 | 0.064 | S | 0.002 | S | 0.19 | I | 0.25 | S | - | - |
| 170 | 2018 | M | 30-34 | 0.38 | I | 0.003 | S | 0.25 | I | 8 | R | - | - |
| 171 | 2018 | M | 25-29 | 2 | R | 0.047 | S | 12 | R | 2 | R | - | - |
| 172 | 2018 | M | 15-19 | 0.25 | I | 0.016 | S | 33 | R | 1 | I | - | - |
| 173 | 2018 | M | 15-19 | 3 | R | 0.003 | S | 1.5 | R | - | - | - | - |
| 174 | 2018 | M | 20-24 | 0.25 | I | 0.006 | S | 1 | R | - | - | - | - |
| 175 | 2018 | M | 15-19 | 0.5 | I | 0.016 | S | 6 | R | - | - | - | - |
| 176 | 2018 | M | 30-34 | 0.094 | I | 0.002 | S | - | - | 4 | R | - | - |
| 177 | 2018 | M | 30-34 | 2 | R | - | - | 4 | R | 0.094 | S | - | - |
| 178 | 2018 | M | 15-19 | 0.19 | I | 0.004 | S | 0.75 | R | 24 | R | - | - |
| 179 | 2018 | M | 15-19 | 0.5 | I | 0.023 | S | 6 | R | 32 | R | - | - |
| 180 | 2018 | M | 15-19 | 0.19 | I | 0.004 | S | 0.75 | R | 24 | R | - | - |
| 181 | 2018 | M | 15-19 | 0.5 | I | 0.023 | S | 6 | R | 32 | R | - | - |
| 182 | 2018 | M | 15-19 | 0.38 | I | 0.008 | S | - | - | 0.75 | I | 0.25 | S |
| 183 | 2018 | M | 40-44 | 0.125 | I | 0.003 | S | 32 | R | 0.75 | I | - | - |
| 184 | 2018 | M | 20-24 | 0.125 | I | 0.002 | S | 0.002 | S | 24 | R | - | - |
| 185 | 2018 | M | 20-24 | 0.064 | S | 0.002 | S | 0.75 | R | - | - | - | - |
|  |  |  |  | MIC | Result | MIC | Result | MIC | Result | MIC | Result | MIC | Result |
| 186 | 2018 | M | 10-14 | 0.125 | I | 0.003 | S | 0.002 | S | 16 | R | 0.19 | S |
| 187 | 2018 | M | 35-39 | 0.064 | S | 0.006 | S | 2 | R | - | - | 0.047 | S |
| 188 | 2018 | M | 25-29 | 0.75 | I | 0.006 | S | 0.75 | R | 0.25 | S | 2 | R |
| 189 | 2018 | F | 30-34 | 0.125 | I | 0.002 | S | 4 | R | 0.5 | I | - | - |
| 190 | 2018 | M | 15-19 | 1.5 | R | 0.023 | S | 4 | R | 1.5 | R | 0.75 | S |
| 191 | 2018 | M | 25-29 | 3 | R | 0.002 | S | 0.25 | I | 4 | R | 0.032 | S |
| 192 | 2018 | M | 15-19 | 0.016 | S | 0.002 | S | 0.5 | I | 2 | R | 0.125 | S |
| 193 | 2018 | F | 35-39 | 0.094 | I | 0.003 | S | 3 | R | 0.125 | S | 0.094 | S |
| 194 | 2018 | M | 15-19 | 0.064 | S | 0.002 | S | 0.004 | S | 0.19 | S | 0.25 | S |
| 195 | 2018 | M | 15-19 | - | - | 0.003 | S | 0.5 | I | 8 | R | 0.064 | S |
| 196 | 2018 | M | 30-34 | - | - | 0.002 | S | 1.5 | R | 16 | R | 0.19 | S |
| 197 | 2018 | M | 55-59 | 0.094 | I | 0.003 | S | 0.75 | R | 0.5 | I | 0.032 | S |
| 198 | 2018 | M | 20-24 | 0.19 | I | 0.004 | S | 0.002 | S | 16 | R | 0.25 | S |
| 199 | 2018 | M | 25-29 | 1.5 | R | 0.016 | S | 0.5 | I | 8 | R | 0.75 | S |
| 200 | 2018 | M | 20-24 | 3 | R | 0.002 | S | 0.75 | R | 0.5 | I | 0.016 | S |
| 201 | 2018 | M | 55-59 | - | - | 0.094 | S | 8 | R | 1.5 | R | 1 | S |
| 202 | 2018 | M | 20-24 | - | - | 0.003 | S | 0.75 | R | 12 | R | 0.064 | S |
| 203 | 2018 | M | 20-24 | 0.064 | S | 0.008 | S | 0.5 | I | 12 | R | 0.047 | S |
| 204 | 2018 | M | 20-24 | - | - | 0.032 | S | 4 | R | 1 | I | 0.5 | S |
| 205 | 2018 | M | 15-19 | - | - | 0.006 | S | 4 | R | 0.75 | I | 0.5 | S |
| 206 | 2018 | M | 30-34 | 0.125 | I | 0.002 | S | 2 | R | 0.19 | S | 0.125 | S |
| 207 | 2018 | M | 55-59 | - | - | 0.032 | S | 4 | R | 0.125 | S | 1 | S |
| 208 | 2018 | M | 15-19 | 0.5 | I | 0.003 | S | 0.38 | I | 24 | R | 0.094 | S |
| 209 | 2018 | M | 40-44 | 0.75 | I | 0.008 | S | 3 | R | 0.5 | I | 0.75 | S |
| 210 | 2018 | M | 15-19 | - | - | 0.004 | S | 1.5 | R | 0.75 | I | 0.023 | S |
| 211 | 2018 | M | 20-24 | 0.38 | I | 0.008 | S* | 0.094 | I | 0.064 | S | 2 | R |
| 212 | 2018 | M | 20-24 | 0.75 | I | 0.023 | S | 2 | R | 0.75 | I | 0.75 | S |
| 213 | 2018 | M | 20-24 | 0.094 | I | 0.002 | S | 0.003 | S | 0.38 | I | 0.125 | S |
| 214 | 2018 | M | 40-44 | 2 | R | 0.016 | S | 0.125 | I | 16 | R | 0.19 | S |
| 215 | 2018 | M | 30-34 | 0.047 | S | 0.002 | S | 0.25 | I | 2 | R | 0.016 | S |
| 216 | 2018 | M | 15-19 | 0.016 | S | 0.002 | S | 0.002 | S | 0.19 | S | 0.064 | S |
| 217 | 2018 | M | 20-24 | 0.38 | I | 0.023 | S | - | - | 1 | I | - | - |
| 218 | 2018 | M | 20-24 | 0.064 | S | 0.002 | S | 0.5 | I | 12 | R | 0.75 | S |
| 219 | 2018 | M | 20-24 | 0.125 | I | 0.008 | S | 1.5 | R | 0.38 | I | 0.25 | S |
| 220 | 2018 | M | 40-44 | 6 | R | 0.003 | S | 0.75 | R | 0.38 | I | 0.064 | S |
| 221 | 2018 | M | 15-19 | 0.094 | I | 0.002 | S | 1 | R | 8 | R | 0.032 | S |
| 222 | 2018 | M | 20-24 | 0.19 | I | 0.006 | S | 0.75 | R | 0.25 | S | 0.19 | S |
| 223 | 2018 | M | 20-24 | 256 | R | 0.003 | S | 6 | R | 16 | R | 0.023 | S |
| 224 | 2018 | M | 15-19 | 0.75 | I | 0.016 | S | 0.125 | I | 0.75 | I | 0.5 | S |
| 225 | 2018 | M | 30-34 | 0.094 | I | 0.002 | S | 1.5 | R | 0.5 | I | 0.25 | S |
| 226 | 2018 | M | 15-19 | 1 | I | 0.064 | S | 0.75 | R | 12 | R | 0.19 | S |
| 227 | 2018 | M | 20-24 | 0.5 | I | 0.002 | S | 0.19 | I | 0.38 | I | 0.032 | S |
| 228 | 2018 | M | 25-29 | 0.064 | S | 0.003 | S | 1.5 | R | 0.25 | S | - | - |
| 229 | 2018 | M | 40-44 | 0.19 | I | 0.003 | S | 1.5 | R | 12 | R | 0.094 | S |
| 230 | 2018 | M | 20-24 | 0.19 | I | 0.012 | S | 0.125 | I | 0.19 | S | 0.19 | S |
| 231 | 2018 | M | 15-19 | 0.19 | I | 0.023 | S | 1.5 | R | 0.19 | S | 0.25 | S |
| 232 | 2018 | M | 25-29 | 0.25 | I | 0.008 | S | 1.5 | R | 0.19 | S | 0.25 | S |
| 233 | 2018 | M | 20-24 | 0.125 | I | 0.002 | S | 0.5 | I | 0.125 | S | 0.047 | S |
| 234 | 2018 | M | 45-49 | 32 | R | 0.006 | S | 1.5 | R | 16 | R | 0.094 | S |
| 235 | 2018 | M | 25-29 | 0.5 | I | 0.023 | S | 4 | R | 0.5 | I | 0.5 | S |
| 236 | 2018 | M | 20-24 | 0.125 | I | 0.003 | S | 0.38 | I | 8 | R | 0.023 | S |
| 237 | 2018 | M | 20-24 | - | - | 0.023 | S | 4 | R | 0.75 | I | 0.38 | S |
| 238 | 2018 | M | 30-34 | - | - | 0.003 | S | 1 | R | 12 | R | 0.047 | S |
| 239 | 2018 | M | 25-29 | - | - | 0.002 | S | 0.003 | S | 0.5 | I | 0.047 | S |
| 240 | 2018 | M | 75-79 | 0.125 | I | 0.006 | S | 6 | R | 0.38 | I | 0.19 | S |
| 241 | 2018 | M | 40-44 | 4 | R | 0.002 | S | 0.5 | I | 6 | R | 0.023 | S |
| 242 | 2018 | M | 30-34 | 16 | R | 0.002 | S | 0.5 | I | 6 | R | 0.047 | S |
| 243 | 2018 | M | 20-24 | 0.19 | I | 0.004 | S | 2 | R | 0.75 | I | 0.25 | S |
| 244 | 2018 | M | 35-39 | 0.125 | I | 0.006 | S | 3 | R | 0.75 | I | 0.125 | S |
| 245 | 2018 | M | 25-29 | 0.064 | S | 0.002 | S | 0.38 | I | 0.25 | S | 0.125 | S |
| 246 | 2018 | M | 15-19 | 4 | R | 0.002 | S | 3 | R | 4 | R | 0.19 | S |
| 247 | 2018 | M | 15-19 | 16 | R | 0.002 | S | 0.002 | S | 0.75 | I | 0.094 | S |
|  |  |  |  | MIC | Result | MIC | Result | MIC | Result | MIC | Result | MIC | Result |
| 248 | 2018 | M | 25-29 | 1 | I | 0.002 | S | 0.5 | I | 16 | R | 0.19 | S |
| 249 | 2018 | M | 20-24 | 0.064 | S | 0.002 | S | 4 | R | 0.25 | S | 0.064 | S |
| 250 | 2018 | F | 40-44 | 24 | R | 0.003 | S | 0.75 | R | 12 | R | 0.094 | S |
| 251 | 2018 | M | 30-34 | 0.5 | I | 0.016 | S | 16 | R | 0.75 | I | 0.094 | S |
| 252 | 2018 | M | 35-39 | 0.094 | I | 0.012 | S | 12 | R | 0.25 | S | 0.094 | S |
| 253 | 2018 | M | 20-24 | 4 | R | 0.002 | S | 0.25 | I | 6 | R | 0.023 | S |
| 254 | 2018 | M | 40-44 | 0.75 | I | 0.003 | S | 0.064 | S | 0.75 | I | 0.094 | S |
| 255 | 2018 | M | 20-24 | 4 | R | 0.002 | S | 0.38 | I | 2 | R | 0.094 | S |
| 256 | 2018 | M | 20-24 | 0.5 | I | 0.012 | S | 8 | R | 0.5 | I | 0.38 | S |
| 257 | 2018 | M | 15-19 | 0.032 | S | 0.002 | S | 0.002 | S | 0.19 | S | 0.016 | S |
| 258 | 2018 | F | 60-64 | 1.5 | R | 0.002 | S | 0.5 | I | 3 | R | 0.125 | S |
| 259 | 2018 | M | 30-34 | 0.016 | S | 0.002 | S | 4 | R | 0.125 | S | 0.125 | S |
| 260 | 2018 | M | 20-24 | 0.19 | I | 0.008 | S | 3 | R | 0.5 | I | 0.094 | S |
| 261 | 2018 | M | 15-19 | 0.064 | S | 0.002 | S | 0.002 | S | 0.25 | S | 0.016 | S |
| 262 | 2019 | M | 20-24 | 4 | R | 0.016 | S | 0.38 | I | 0.125 | S | 0.047 | S |
| 263 | 2019 | M | 20-24 | - | - | 0.016 | S | 0.38 | I | 3 | R | 0.032 | S |
| 264 | 2019 | M | 15-19 | - | - | 0.016 | S | 0.25 | I | 4 | R | 0.064 | S |
| 265 | 2019 | M | 20-24 | 0.75 | I | 0.016 | S | 0.002 | S | 0.38 | I | 0.064 | S |
| 266 | 2019 | M | 20-24 | 0.38 | I | 0.016 | S | 3 | R | 0.75 | I | 0.5 | S |
| 267 | 2019 | M | 25-29 | 8 | R | 0.016 | S | 0.25 | I | 12 | R | 0.064 | S |
| 268 | 2019 | M | 20-24 | 0.19 | I | 0.016 | S | 0.004 | S | 0.5 | I | 1.5 | R |
| 269 | 2019 | M | 20-24 | 1 | I | 0.016 | S | 1 | R | 8 | R | 0.047 | S |
| 270 | 2019 | M | 20-24 | - | - | 0.016 | S | - | - | 4 | R | 0.094 | S |
| 271 | 2019 | F | 25-29 | 0.094 | I | 0.016 | S | 1 | R | 4 | R | 0.016 | S |
| 272 | 2019 | M | 25-29 | - | - | 0.023 | S | - | - | 1 | I | 0.25 | S |
| 273 | 2019 | M | 15-19 | - | - | 0.016 | S | - | - | 0.25 | S | 0.047 | S |
| 274 | 2019 | M | 35-39 | - | - | 0.023 | S | 4 | R | 0.5 | I | 0.25 | S |
| 275 | 2019 | M | 20-24 | - | - | 0.032 | S | - | - | 0.75 | I | 0.032 | S |
| 276 | 2019 | M | 15-19 | 0.047 | S | 0.016 | S | 0.002 | S | 12 | R | 0.125 | S |
| 277 | 2019 | M | 30-34 | - | - | 0.016 | S | - | - | 0.125 | S | 0.094 | S |
| 278 | 2019 | M | 20-24 | - | - | 0.016 | S | 16 | R | 8 | R | 0.032 | S |
| 279 | 2019 | M | 15-19 | 0.75 | I | 0.016 | S | 0.002 | S | 0.25 | S | 0.094 | S |
| 280 | 2019 | M | 25-29 | 0.094 | I | 0.016 | S | 1 | R | 1 | I | 0.023 | S |
| 281 | 2019 | M | 25-29 | - | - | 0.016 | S | - | - | 0.25 | S | 0.094 | S |
| 282 | 2019 | M | 35-39 | 2 | R | 0.016 | S | 0.38 | I | 0.19 | S | 0.064 | S |
| 283 | 2019 | M | 15-19 | 16 | R | 0.016 | S | 0.38 | I | 24 | R | 0.032 | S |
| 284 | 2019 | M | 25-29 | - | - | 0.016 | S | 1.5 | R | 6 | R | 0.047 | S |
| 285 | 2019 | M | 20-24 | - | - | 0.016 | S | - | - | 0.5 | I | 0.125 | S |
| 286 | 2019 | M | 25-29 | 0.125 | I | - | - | - | - | 8 | R | 0.016 | S |
| 287 | 2019 | M | 40-44 | 3 | R | - | - | - | - | 4 | R | 0.016 | S |
| 288 | 2019 | M | 25-29 | 0.75 | I | 0.023 | S | 0.023 | S | 0.5 | I | 0.064 | S |
| 289 | 2019 | M | 20-24 | 16 | R | 0.023 | S | 0.75 | R | 12 | R | 0.023 | S |
| 290 | 2019 | M | 20-24 | 0.25 | I | 0.008 | S | 16 | R | 0.75 | I | 0.25 | S |
| 291 | 2019 | M | 15-19 | 0.047 | S | 0.016 | S | 0.002 | S | 3 | R | 0.023 | S |
| 292 | 2019 | M | 20-24 | 0.064 | S | - | - | - | - | 0.25 | S | 0.016 | S |
| 293 | 2019 | M | 15-19 | 0.023 | S | - | - | - | - | 0.032 | S | 0.064 | S |
| 294 | 2019 | M | 15-19 | - | - | - | - | 1.5 | R | 12 | R | 0.25 | S |
| 295 | 2019 | M | 30-34 | 0.5 | I | - | - | 2 | R | 0.75 | I | 0.5 | S |
| 296 | 2019 | M | 30-34 | 0.25 | I | - | - | 1.5 | R | 0.38 | I | 0.25 | S |
| 297 | 2019 | M | 30-34 | 6 | R | - | - | 0.5 | I | 16 | R | 0.032 | S |
| 298 | 2019 | M | 25-29 | - | - | - | - | 1 | R | 6 | R | 0.125 | S |
| 299 | 2019 | M | 20-24 | 0.19 | I | 0.012 | S | 32 | R | - | - | 0.5 | S |
| 300 | 2019 | M | 25-29 | 0.25 | I | 0.25 | S | 4 | R | 0.5 | I | 0.38 | S |
| 301 | 2019 | M | 20-24 | 0.094 | I | 0.002 | S | - | - | - | - | 0.125 | S |
| 302 | 2019 | F | 35-39 | 0.75 | I | - | - | 6 | R | 1.5 | R | 0.5 | S |
| 303 | 2019 | M | 35-39 | - | - | - | - | 0.75 | R | 12 | R | 0.125 | S |
| 304 | 2019 | M | 35-39 | - | - | - | - | 3 | R | 0.25 | S | 0.25 | S |
| 305 | 2019 | M | 20-24 | 0.25 | I | 0.006 | S | - | - | - | - | 0.75 | S |
| 306 | 2019 | M | 20-24 | - | - | - | - | 0.004 | S | 0.25 | S | 8 | R |
| 307 | 2019 | M | 20-24 | 256 | R | 0.008 | S | - | - | - | - | 0.75 | S |
| 308 | 2019 | M | 20-24 | 0.19 | I | 0.006 | S | - | - | 0.5 | I | 0.75 | S |
| 309 | 2019 | M | 20-24 | 0.064 | S | 0.002 | S | - | - | - | - | 0.064 | S |
| 310 | 2019 | M | 25-29 | 0.064 | S | 0.012 | S | - | - | 0.5 | I | 0.75 | S |
|  |  |  |  | MIC | Result | MIC | Result | MIC | Result | MIC | Result | MIC | Result |
| 311 | 2019 | M | 25-29 | 0.094 | I | 0.002 | S | - | - | 16 | R | 0.19 | S |
| 312 | 2019 | M | 25-29 | 0.19 | I | 0.032 | S | - | - | - | - | 256 | R |
| 313 | 2019 | M | 30-34 | 256 | R | 0.002 | S | - | - | - | - | 0.125 | S |
| 314 | 2019 | M | 15-19 | 0.094 | I | 0.002 | S | - | - | - | - | 0.38 | S |
| 315 | 2019 | M | 20-24 | 0.064 | S | 0.002 | S | - | - | - | - | 0.5 | S |
| 316 | 2019 | M | 15-19 | 0.023 | S | 0.002 | S | - | - | - | - | 0.064 | S |
| 317 | 2019 | M | 15-19 | 0.016 | S | 0.002 | S | - | - | - | - | 0.016 | S |
| 318 | 2019 | M | 25-29 | 3 | R | 0.064 | S | - | - | - | - | 0.38 | S |
| 319 | 2019 | M | 15-19 | 0.25 | I | 0.004 | S | - | - | - | - | 0.38 | S |
| 320 | 2019 | M | 25-29 | 0.094 | I | 0.006 | S | - | - | - | - | 0.38 | S |
| 321 | 2019 | M | 15-19 | 256 | R | 0.003 | S | - | - | - | - | 0.38 | S |
| 322 | 2019 | M | 35-39 | 0.19 | I | 0.002 | S | - | - | - | - | 0.38 | S |
| 323 | 2019 | F | 25-29 | 0.064 | S | 0.002 | S | - | - | - | - | 0.19 | S |
| 324 | 2019 | F | 35-39 | 8 | R | 0.002 | S | - | - | - | - | 0.125 | S |
| 325 | 2019 | M | 20-24 | 0.064 | S | 0.016 | S | - | - | - | - | 0.25 | S |
| 326 | 2019 | M | 20-24 | 0.125 | I | 0.004 | S | - | - | - | - | 0.25 | S |
| 327 | 2019 | M | 20-24 | 0.125 | I | 0.002 | S | - | - | - | - | 0.19 | S |
| 328 | 2019 | M | 20-24 | 24 | R | 0.002 | S | - | - | - | - | 0.125 | S |
| 329 | 2019 | M | 20-24 | 0.094 | I | 0.023 | S | - | - | - | - | 0.125 | S |
| 330 | 2019 | M | 35-39 | 0.125 | I | 0.002 | S | - | - | - | - | 0.125 | S |
| 331 | 2019 | M | 30-34 | 0.38 | I | 0.016 | S | - | - | - | - | 0.75 | S |
| 332 | 2019 | M | 25-29 | 256 | R | 0.002 | S | 0.75 | R | - | - | 0.125 | S |
| 333 | 2019 | M | 30-34 | 1 | I | 0.023 | S | 12 | R | - | - | 2 | R |
| 334 | 2019 | M | 25-29 | 0.25 | I | 0.012 | S | - | - | - | - | 0.75 | S |
| 335 | 2019 | F | 20-24 | 32 | R | 0.002 | S | - | - | - | - | - | - |
| 336 | 2019 | M | 20-24 | 3 | R | 0.002 | S | - | - | - | - | 0.047 | S |
| 337 | 2019 | M | 20-24 | 0.19 | I | 0.006 | S | 1.5 | R | - | - | 0.5 | S |
| 338 | 2019 | M | 20-24 | 0.25 | I | 0.008 | S | - | - | - | - | 0.75 | S |
| 339 | 2019 | M | 20-24 | 48 | R | 0.002 | S | 1 | R | - | - | 0.125 | S |
| 340 | 2019 | M | 25-29 | 1 | I | 0.002 | S | 0.5 | I | - | - | 0.19 | S |
| 341 | 2019 | M | 30-34 | 0.5 | I | 0.002 | S | 0.75 | R | - | - | 0.064 | S |
| 342 | 2019 | M | 20-24 | 0.19 | I | 0.004 | S | 1.5 | R | - | - | 0.38 | S |
| 343 | 2019 | M | 15-19 | 6 | R | 0.002 | S | 0.75 | R | - | - | 0.094 | S |
| 344 | 2019 | M | 20-24 | 0.25 | I | 0.016 | S | 1.5 | R | - | - | 0.5 | S |
| 345 | 2019 | M | 25-29 | 0.064 | S | 0.004 | S | 6 | R | - | - | 0.094 | S |
| 346 | 2019 | M | 25-29 | 3 | R | 0.002 | S | 0.25 | I | - | - | 0.094 | S |
| 347 | 2019 | M | 40-44 | 4 | R | 0.004 | S | 0.75 | R | - | - | 0.38 | S |
| 348 | 2019 | M | 20-24 | 0.75 | I | 0.006 | S | - | - | - | - | 256 | R |
| 349 | 2019 | M | 25-29 | 6 | R | 0.002 | S | 0.38 | I | - | - | 0.125 | S |
| 350 | 2019 | M | 25-29 | 0.38 | I | 0.002 | S | 0.064 | S | - | - | 0.094 | S |
| 351 | 2019 | M | 20-24 | 8 | R | 0.002 | S | 0.75 | R | - | - | 0.125 | S |
| 352 | 2019 | M | 25-29 | 0.75 | I | 0.002 | S | 4 | R | - | - | - | - |
| 353 | 2019 | M | 30-34 | 0.19 | I | 0.004 | S | 1.5 | R | - | - | 0.38 | S |
| 354 | 2019 | M | 30-34 | 0.094 | I | 0.002 | S | 1 | R | - | - | 0.125 | S |
| 355 | 2019 | M | 35-39 | 0.5 | I | 0.012 | S | 0.75 | R | - | - | 0.38 | S |
| 356 | 2019 | M | 60-64 | 24 | R | 0.004 | S | 1.5 | R | 24 | R | 0.19 | S |
| 357 | 2019 | F | 30-34 | 0.75 | I | 0.002 | S | 0.25 | I | - | - | 0.5 | S |
| 358 | 2019 | M | 30-34 | 0.094 | I | 0.002 | S | 0.19 | I | - | - | 0.19 | S |
| 359 | 2019 | M | 15-19 | 0.25 | I | 0.002 | S | 0.008 | S | - | - | 3 | R |
| 360 | 2019 | M | 20-24 | 4 | R | 0.003 | S | 0.75 | R | 0.5 | I | 0.19 | S |
| 361 | 2019 | M | 20-24 | 3 | R | 0.003 | S | 1 | R | 6 | R | 0.25 | S |
| 362 | 2019 | M | 25-29 | 0.75 | I | 0.008 | S | 1 | R | 0.75 | I | 1 | S |
| 363 | 2019 | M | 35-39 | 3 | R | 0.004 | S | 2 | R | 16 | R | 0.38 | S |
| 364 | 2019 | M | 15-19 | 0.016 | S | 0.002 | S | 1 | R | 0.19 | S | 0.38 | S |
| 365 | 2019 | M | 25-29 | 0.38 | I | 0.012 | S | 3 | R | 1 | I | 1 | S |
| 366 | 2019 | M | 20-24 | 6 | R | 0.002 | S | 0.25 | I | 0.5 | I | 0.25 | S |
| 367 | 2019 | M | 20-24 | 1 | I | 0.002 | S | 0.023 | S | 0.125 | S | 0.016 | S |
| 368 | 2019 | M | 20-24 | 8 | R | 0.002 | S | 0.25 | I | 0.5 | I | 0.19 | S |
| 369 | 2019 | M | 20-24 | 0.125 | I | 0.008 | S | 2 | R | 0.75 | I | 0.5 | S |
| 370 | 2019 | M | 30-34 | 0.75 | I | 0.002 | S | 0.25 | I | 24 | R | 0.19 | S |
| 371 | 2019 | M | 15-19 | 0.75 | I | 0.002 | S | 1 | R | 16 | R | 0.25 | S |
| 372 | 2019 | M | 20-24 | 2 | R | 0.003 | S | 0.25 | I | 32 | R | 0.38 | S |
| 373 | 2019 | M | 15-19 | 0.25 | I | 0.012 | S | 1.5 | R | 0.75 | I | 0.5 | S |
|  |  |  |  | MIC | Result | MIC | Result | MIC | Result | MIC | Result | MIC | Result |
| 374 | 2019 | M | 20-24 | 0.094 | I | 0.002 | S | 0.125 | I | 4 | R | 0.19 | S |
| 375 | 2019 | M | 35-39 | 0.016 | S | 0.002 | S | 3 | R | 0.25 | S | 1 | S |
| 376 | 2019 | M | 20-24 | 0.094 | I | 0.002 | S | 0.5 | I | 8 | R | 0.125 | S |
| 377 | 2019 | M | 25-29 | 12 | R | 0.002 | S | 1 | R | 0.75 | I | 0.125 | S |
| 378 | 2019 | M | 40-44 | 0.016 | S | 0.002 | S | 2 | R | 0.38 | I | 0.75 | S |
| 379 | 2019 | M | 30-34 | 2 | R | 0.002 | S | 1 | R | 32 | R | 0.19 | S |
| 380 | 2019 | M | 20-24 | 0.064 | S | 0.002 | S | 0.002 | S | 0.5 | I | 0.5 | S |
| 381 | 2019 | M | 50-54 | 0.19 | I | 0.012 | S | 4 | R | 0.75 | I | 0.75 | S |
| 382 | 2019 | M | 25-29 | 0.125 | I | 0.002 | S | 1 | R | 16 | R | 0.25 | S |
| 383 | 2019 | M | 15-19 | 48 | R | 0.003 | S | 1.5 | R | 24 | R | 0.38 | S |
| 384 | 2019 | M | 35-39 | 256 | R | 0.003 | S | 1 | R | 1 | I | 0.25 | S |
| 385 | 2019 | M | 15-19 | 12 | R | 0.004 | S | 3 | R | 24 | R | 0.5 | S |
| 386 | 2019 | M | 40-44 | 0.19 | I | 0.004 | S | 0.5 | I | 16 | R | 1 | S |
| 387 | 2019 | M | 20-24 | 1.5 | R | 0.002 | S | 0.5 | I | 12 | R | 0.5 | S |
| 388 | 2020 | M | 20-24 | 0.25 | I | 0.003 | S | - | - | 0.38 | I | 0.75 | S |
| 389 | 2020 | M | 35-39 | 0.047 | S | 0.002 | S | 0.002 | S | 0.125 | S | 0.094 | S |
| 390 | 2020 | M | 20-24 | 0.5 | I | 0.012 | S | - | - | 0.75 | I | 1 | S |
| 391 | 2020 | M | 30-34 | 0.38 | I | 0.003 | S | - | - | 12 | R | 0.5 | S |
| 392 | 2020 | M | 25-29 | 32 | R | 0.002 | S | 0.25 | I | 8 | R | 0.064 | S |
| 393 | 2020 | M | 15-19 | 32 | R | 0.002 | S | 0.5 | I | 8 | R | 0.25 | S |
| 394 | 2020 | M | 30-34 | 0.125 | I | 0.002 | S | 0.5 | I | 0.38 | I | 0.25 | S |
| 395 | 2020 | M | 20-24 | 0.19 | I | 0.003 | S | 0.006 | S | 0.38 | I | 2 | R |
| 396 | 2020 | M | 20-24 | 32 | R | 0.004 | S | - | - | 16 | R | 0.38 | S |
| 397 | 2020 | M | 30-34 | 0.125 | I | 0.002 | S | 0.25 | I | 8 | R | 0.5 | S |
| 398 | 2020 | M | 30-34 | 0.5 | I | 0.002 | S | - | - | 12 | R | 0.19 | S |
| 399 | 2020 | M | 20-24 | 0.38 | I | 0.006 | S | 1.5 | R | 0.5 | I | 1 | S |
| 400 | 2020 | M | 30-34 | 2 | R | 0.003 | S | 0.25 | I | 2 | R | 0.094 | S |
| 401 | 2020 | M | 15-19 | 0.25 | I | 0.016 | S | 2 | R | 1 | I | 1 | S |
| 402 | 2020 | M | 40-44 | 0.125 | I | 0.008 | S | 0.002 | S | 0.125 | S | 0.75 | S |
| 403 | 2020 | M | 40-44 | 0.016 | S | 0.002 | S | 0.002 | S | 0.25 | S | 0.75 | S |
| 404 | 2020 | M | 15-19 | 0.75 | I | 0.002 | S | 0.5 | I | 16 | R | 0.25 | S |
| 405 | 2020 | M | 25-29 | 0.016 | S | 0.002 | S | 0.002 | S | 0.094 | S | 0.094 | S |
| 406 | 2020 | M | 20-24 | 0.5 | I | 0.003 | S | 1 | R | 1.5 | R | 0.25 | S |
| 407 | 2020 | M | 50-54 | 0.25 | I | 0.006 | S | 0.125 | I | 4 | R | 0.5 | S |
| 408 | 2020 | M | 20-24 | 3 | R | 0.003 | S | 1 | R | 12 | R | 0.25 | S |
| 409 | 2020 | M | 35-39 | 6 | R | 0.002 | S | 0.75 | R | 0.75 | I | 0.094 | S |
| 410 | 2020 | M | 40-44 | 0.125 | I | 0.002 | S | 0.75 | R | 0.38 | I | 0.25 | S |
| 411 | 2020 | M | 35-39 | 0.125 | I | 0.002 | S | 0.004 | S | 0.75 | I | 1 | S |
| 412 | 2020 | M | 20-24 | 0.016 | S | 0.002 | S | 0.002 | S | 0.5 | I | 0.5 | S |
| 413 | 2020 | M | 20-24 | 1 | I | 0.19 | S | 6 | R | 0.5 | I | 256 | R |
| 414 | 2020 | M | 25-29 | 0.38 | I | 0.008 | S | 4 | R | 0.5 | I | 0.75 | S |
| 415 | 2020 | M | 20-24 | 0.25 | I | 0.012 | S | 3 | R | 1 | I | 1 | S |
| 416 | 2020 | M | 25-29 | 0.047 | S | 0.012 | S | 3 | R | 0.25 | S | 0.125 | S |
| 417 | 2020 | M | 25-29 | 6 | R | 0.002 | S | 0.5 | I | 0.5 | I | 0.047 | S |
| 418 | 2020 | M | 15-19 | 0.25 | I | 0.002 | S | 0.19 | I | 16 | R | 0.064 | S |
| 419 | 2020 | M | 20-24 | 0.064 | S | 0.002 | S | 0.75 | R | 0.25 | S | 0.19 | S |
| 420 | 2020 | M | 45-49 | 0.064 | S | 0.002 | S | 0.023 | S | 2 | R | 0.125 | S |
| 421 | 2020 | M | 20-24 | 0.094 | I | 0.003 | S | 1 | R | 3 | R | 0.047 | S |
| 422 | 2020 | M | 25-29 | 32 | R | 0.032 | S | 3 | R | 16 | R | 0.094 | S |
| 423 | 2020 | M | 55-59 | 2 | R | 0.002 | S | 0.5 | I | 0.25 | S | 0.016 | S |
| 424 | 2020 | M | 30-34 | 0.125 | I | 0.004 | S | 6 | R | 0.25 | S | 0.064 | S |
| 425 | 2020 | M | 25-29 | 12 | R | 0.004 | S | 1 | R | 12 | R | 0.125 | S |
| 426 | 2020 | M | 35-39 | 1 | I | 0.006 | S | 0.5 | I | 0.25 | S | 0.125 | S |
| 427 | 2020 | M | 20-24 | 0.064 | S | 0.002 | S | 0.5 | I | 0.5 | I | 0.19 | S |
| 428 | 2020 | M | 45-49 | 3 | R | 0.003 | S | 1.5 | R | 6 | R | 0.125 | S |
| 429 | 2020 | M | 30-34 | 0.064 | S | 0.002 | S | 2 | R | 0.25 | S | 0.064 | S |
| 430 | 2020 | M | 25-29 | 2 | R | 0.002 | S | 0.25 | I | 0.032 | S | 0.032 | S |
| 431 | 2020 | M | 20-24 | 4 | R | 0.008 | S | 0.38 | I | 6 | R | 0.25 | S |
| 432 | 2020 | M | 30-34 | 0.75 | I | 0.002 | S | 0.25 | I | 8 | R | 0.125 | S |
| 433 | 2020 | M | 25-29 | 0.125 | I | 0.004 | S | 2 | R | 0.5 | I | 0.25 | S |

F, female; I, intermediate; M, male; -, not tested; R, resistance; S, susceptibility.
\*The specimen was not viable for confirmatory Etest. Accordingly, the susceptibility characterisation was based on whole-genome sequencing and a lack of all known genetic resistance determinants for ceftriaxone in the genome sequence.

